# Safety and immunogenicity following co-administration of Yellow fever vaccine with Tick-borne encephalitis or Japanese encephalitis vaccines: Results from an open label, non-randomized clinical trial

**DOI:** 10.1101/2022.07.05.22277040

**Authors:** John Tyler Sandberg, Marie Löfling, Renata Varnaitė, Johanna Emgård, Nabil Al-Tawil, Lars Lindquist, Sara Gredmark-Russ, Jonas Klingström, Karin Loré, Kim Blom, Hans-Gustaf Ljunggren

## Abstract

**Background:** Flavivirus infections pose a significant global health burden underscoring the need for the development of safe and efficient vaccination strategies. Available flavivirus vaccines are from time to time concomitantly delivered to individuals in need. Co-administration of different vaccines saves time and visits to health care units and vaccine clinics. It serves to provide protection against multiple pathogens in a shorter time-span, both for individuals living in, or travelling to, endemic areas. However, safety and immunogenicity-related responses have not been appropriately evaluated upon concomitant delivery of these vaccines. Therefore, we performed an open label, non-randomized clinical trial studying the safety and immunogenicity following concomitant delivery of the Yellow fever virus (YFV) vaccine with Tick-borne encephalitis virus (TBEV) and Japanese encephalitis virus (JE) virus vaccines.

**Methods and findings:** Following screening, healthy study subjects were enrolled into different cohorts receiving either TBEV and YFV vaccines, JEV and YFV vaccines, or in control groups receiving only the TBEV, JEV, or YFV vaccine. Concomitant delivery was given in the same or different upper arms for comparison in the co-vaccination cohorts. Adverse effects were recorded throughout the study period and blood samples were taken before and at multiple time-points following vaccination to evaluate immunological responses to the vaccines. Adverse events were predominantly mild in the study groups. Four serious adverse events (SAE) were reported throughout the trial, none of them deemed related to vaccination. The development of neutralizing antibodies (nAbs) against TBEV, JEV, or YFV was not affected by the concomitant vaccination strategy. Concomitant vaccination in the same or different upper arms did not significantly affect safety or immunogenicity-related outcomes. Exploratory studies on immunological effects were additionally performed and included studies of lymphocyte activation, correlates associated with germinal center activation, and plasmablast expansion.

**Conclusions:** Inactivated TBEV or JEV vaccines can be co-administered with the live attenuated YFV vaccine without an increased risk of adverse events and without reduced development of nAbs to the respective viruses. The vaccines can be delivered in the same upper arm without negative outcome.

**Trial registration:** Eudra CT 2017-002137-32

**Author summary:** *Why was this study done?:*

- Flavivirus infections pose a significant global health burden, underscoring the need for the development of safe and efficient vaccination strategies
- Co-administration of different flavivirus vaccines saves time and visits to health care units and vaccine clinics. It serves to provide protection against multiple pathogens in a shorter time-span, both for individuals living in or travelling to endemic areas
- Safety and immunogenicity-related responses have not been appropriately evaluated upon co-administration of many vaccines including currently used flavivirus vaccines, such as Yellow fever virus (YFV), Tick-borne encephalitis virus (TBEV), and Japanese encephalitis virus (JE) virus vaccines
- Because of this, we performed an open label, non-randomized clinical trial studying the safety and immunogenicity following co-administration of YFV vaccine with TBEV and JEV vaccines

*What did the researchers find?:*

- Adverse events, neutralizing antibodies (nAbs) and other related immunological parameters were not adversely affected by concomitant delivery of the vaccines
- Concomitant vaccination in the same versus different upper arms of study participants did not significantly affect safety or immunogenicity outcomes

*What do these findings mean?:*

- Co-administration of YFV vaccine and TBEV or JEV vaccines is feasible without increased risk of adverse events or reduced development of nAbs against the respective viruses. Furthermore, the vaccines can safely be delivered in the same upper arm without negative outcome

## Introduction

Yellow fever virus (YFV), Tick-borne encephalitis virus (TBEV), and Japanese encephalitis virus (JEV) all belong to the *Flavivirus* genus of the *Flaviviridae* family of viruses. These viruses pose major global health challenges, both for inhabitants in affected areas (around 1/3 of the world’s population) as well as for travelers to these areas [1–5]. Infection with these viruses is associated with significant morbidities, hospitalization costs, sick leave-associated costs, and loss of productivity as well as over 100,000 of fatal outcomes per year [2,4,6–9]. No specific treatment or antiviral drugs for these diseases exists, though development of antiviral drug candidates has been initiated [10–14]. There are, however, approved vaccines against these flaviviruses [15–17]. These vaccines are frequently used to prevent disease among inhabitants living in endemic areas and for travelers visiting these areas.

The vaccine regimen and outcome of vaccinations against YFV, TBEV, and JEV infections differ significantly. After a single dose, the live attenuated YFV vaccine provides at least 10 years, and possibly life-long, immunity [18–20]. In contrast, the current inactivated TBEV and JEV vaccines licensed in Europe require multiple doses for primary immunization and regular booster doses to maintain immunity [15,16,20,21].

Safe and efficient vaccine administration strategies are of significant importance [22]. Rigorous clinical trials have assessed the safety and immunogenicity of most currently approved vaccines. However, data are limited with respect to interactions of many vaccines, including the YFV vaccine, with other vaccines including all currently available flavivirus vaccines [23–32]. Published studies are limited to a few reports on concomitant vaccination of the YFV vaccine with other vaccines including smallpox, Bacillus Calmett-Guérin (BCG), hepatitis B or cholera vaccines [33–35].

Endemic flaviviruses overlap in some parts of the world, affecting local inhabitants as well as travelers with needs for protection. Simultaneous vaccination with different vaccines saves time and visits to health care units and vaccine clinics. It provides protection against multiple pathogens in a shorter time-span when an individual may be vulnerable to infection. As mentioned, there is also often a need for individuals to rapidly protect themselves from flaviviruses prior to travelling to endemic areas. This has been further emphasized more recently given that present flavivirus endemic areas are currently increasing, likely due to changes in climate change and other factors [1,36–38]. Together, this has called for more systematic evaluations of the effects of concomitant delivery of flavivirus vaccines, particularly with respect to safety and immunogenicity.

To this end, we carried out an open label, non-randomized clinical trial assessing the safety and immunogenicity of concomitant vaccination with different commonly used flavivirus vaccines. The clinical trial included healthy adult volunteers who were concomitantly vaccinated with TBEV and YFV vaccines, JEV and YFV vaccines, or with only one of the three respective vaccines. Half of the concomitantly vaccinated study participants received both vaccines in the same upper arm while the other half received the vaccines in different upper arms. Blood samples were taken before and at multiple time-points following vaccinations. Safety and immunological responses including nAbs were evaluated. Collectively, the study provides safety and immunogenicity outcomes following concomitant vaccination with different types of flavivirus vaccines.

## Methods

### Ethical and regulatory approval

The study was approved by the Stockholm Local Regional Ethical Committee (2017/1433-31/1) and the Swedish Medical Products Agency (5.1-2017-52376). It is registered in the European database (Eudra CT 2017-002137-32). All study volunteers signed informed consent documents in line with the ethical approval and clinical trial protocol.

### Study design and participants

An open label, non-randomized academic (non-Pharma sponsored) clinical trial was conducted in order to assess safety and immunological responses of concomitant vaccination with three currently licensed flavivirus vaccines. The trial was conducted at the Karolinska University Hospital, Stockholm, Sweden. An independent data monitoring process was setup by the Karolinska Trial Alliance (KTA) support organization (KTA Support) in order to review safety data and the progress of the study according to the clinical trial protocol. The monitoring process also included review of the trial’s conductance in accordance with principles of good clinical practice (ICH-GCP)[39]. Volunteer recruitment, medical examination, vaccination, and peripheral blood sampling were handled by the KTA Phase I unit at the Karolinska University Hospital. Recording of informed consent, adherence to set inclusion and exclusions criteria including previous vaccinations, age, gender, and a standardized medical examination was performed (including temperature, pulse, blood pressure, and pregnancy test) prior to inclusion in the study. Inclusion criteria allowed volunteers between 18 to 55 years of age who wanted protection to YFV, TBEV, and/or JEV, and who were willing to provide written consent to enter the study. Exclusion criteria included, but were not limited to, previous known infection with or vaccination against YFV, TBEV, and JEV, allergy to eggs, poultry or fructose, immunocompromising diseases, autoimmune diseases, medication for cancer or immunosuppression, HIV or HCV infection, fever within one week of scheduled vaccination, pregnancy, hemophilia, and/or involvement in other medical studies. All study volunteers were properly insured with respect to vaccination and blood sampling risks. Study participants were entitled to leave the study at any time without reason. Further details with respect to exclusion criteria and specific criteria for termination of the study are outlined in the clinical trial protocol.

### Vaccines

The following vaccines were used in the trial: Stamaril^®^ (Sanofi), live, attenuated YFV 17D strain produced in pathogen-free chick embryo cells, 0.5 ml, not less than 1,000 IU. The vaccine was provided in freeze dried powder form and reconstituted with provided saline solution and was administered subcutaneously. IXIARO^®^ (Valneva), inactivated, alum-adjuvanted, Vero cell-derived vaccine based on JEV strain SA_14_-14-2, 0.5 ml. The vaccine was provided in the form of pre-filled syringe attached without needle containing an 0.5 mL dose and was administered intramuscularly according to the manufacturer’s label. FSME IMMUN^®^ (Pfizer), inactivated, alum-adjuvanted, chick embryo cell derived vaccine based on the Neudörfl strain, 0.5 ml. The vaccine was provided as a pre-filled syringe attached without needle containing a 0.5 mL dose and was administered intramuscularly according to the manufacturer’s label. All vaccines were obtained from Apoteket AB, Karolinska University Hospital, Solna, Sweden and kept at the Phase-1 unit, KTA, Karolinska University Hospital, at its Huddinge site.

### Study design and cohorts

The study was initially designed to include a total of 140 healthy volunteers. Forty study participants were to receive both TBEV and YFV vaccines (cohort A). Twenty of these participants were to receive the vaccines in different upper arms (sub-cohort A1) and the other 20 in the same upper arm (sub-cohort A2). The next 40 study participants were to receive both JEV and YFV vaccines (cohort B). Similarly, 20 of these participants were to receive the vaccines in different upper arms (sub-cohort B1) and the other 20 in the same upper arm (sub-cohort B2). The remaining three cohorts of 20 study participants per cohort, were aimed to serve as study controls and were to receive either, the TBEV vaccine (cohort C), JEV vaccine (cohort D), or YFV vaccine (cohort E).

Upon initiation of the clinical trial, a total of 161 healthy volunteers were screened for enrollment (**Fig 1A**). 145 study participants were found eligible for entering the study and assigned to it. Of these, 43 participants were assigned to cohort A (A1, 23 participants; A2, 20 participants), 42 participants to cohort B (B1, 21 participants; B2, 21 participants), and 20 participants each to cohorts C, D, and E. Cohort A1 had in total three study participant dropouts and cohorts B1, B2, and C had one study participant dropout each, following enrollment to the study. Hence, 139 study subjects in total completed the trial. A total of 13 missed visits out of 1,150 total planned visits occurred during the course of the trial.

**Fig. 1.**
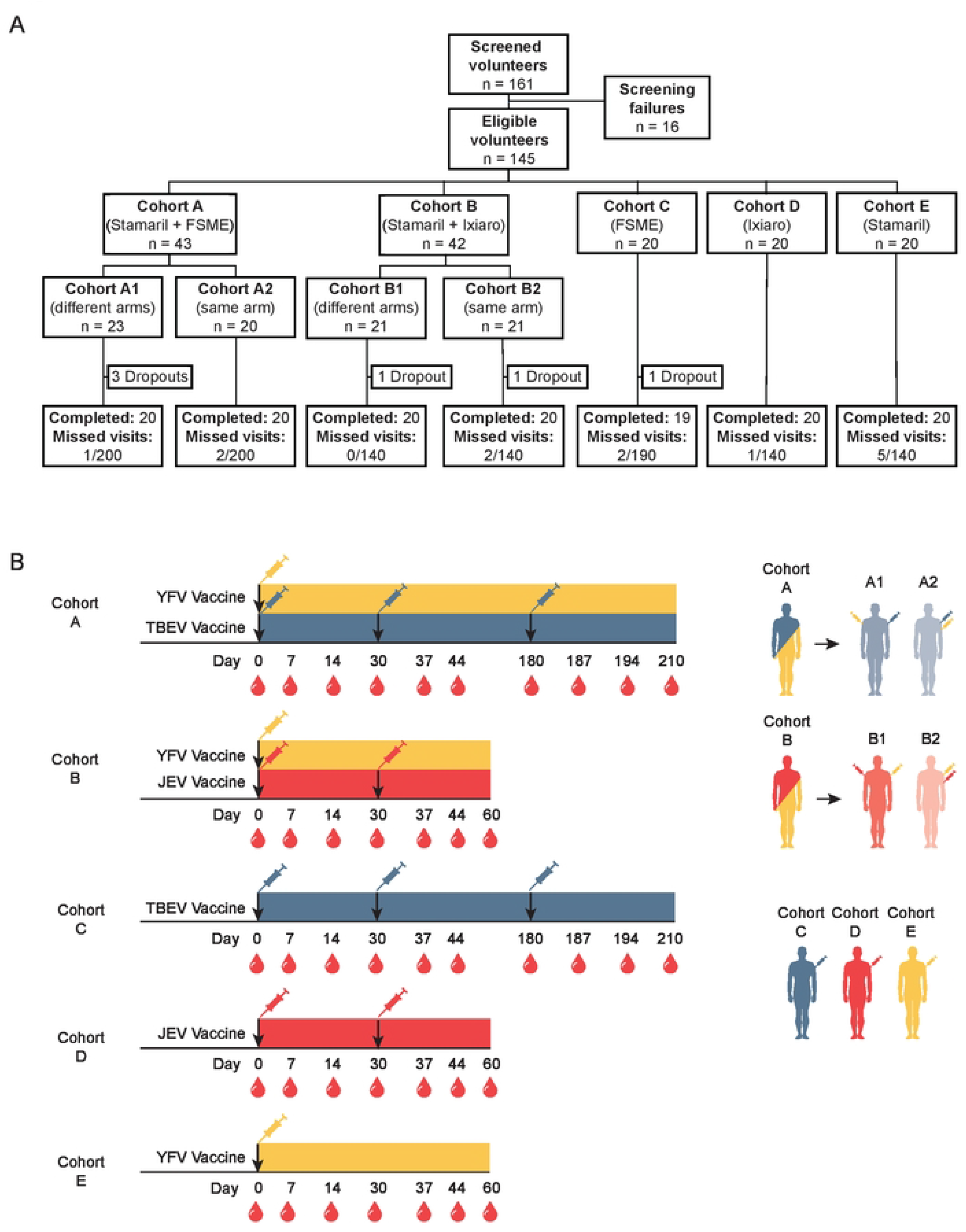
Clinical trial layout for study participants and vaccination strategy with sampling timeline. (**A)** The flowchart depicts the screening and enrollment of healthy volunteers in the study. Enrolled study participants were divided into seven vaccine cohorts. Study participant dropouts and total missed visits are depicted. **(B)** Schematic of vaccination strategy and sampling schedule for enrolled study participants. Peripheral blood and serum samples were taken at the time-points indicated by blood drop symbol. See Material and Methods for vaccination details.

### Procedures

Vaccinations were given in accordance with the clinical trial protocol and good clinical practice (GCP), with intervals for primary vaccinations as recommended in FASS (Pharmaceutical Specialties in Sweden): FSME IMMUN, three doses 0-, 1-and 5-month intervals; IXIARO, two doses, 0- and 1-month interval; Stamaril, one dose. For cohorts A1, A2, B1, B2, C, and D, blood and serum were sampled before each vaccination, and then at day 7 (+/- 1 day) and at day 14. Blood and serum were also sampled 30 days after the last vaccination (+14 days). For cohort E, blood and serum were sampled before the vaccination, and then at day 7 (+/- 1 day), at day 14, at day 30 (+/-2 days), day 37 (+/- 1 day), day 44 (+5 days) and day 60 (+14 days). This schedule led to the following approximate time allocations in terms of days for blood and serum samplings: cohorts A and C, days 0 (first vaccination), 7, 14, 30 (second vaccination), 37, 44, 180 (third vaccination), 187, 194, and 210; cohorts B and D, days 0 (first vaccination), 7, 14, 30 (second vaccination), 37, 44, and 60; cohort E, days 0 (vaccination), 7, 14, 30, 37, 44, and 60 (**Fig 1B**). At each sampling time-point, 40-45 ml of venous blood from study participants was collected in vacuum serum and EDTA tubes (BD). PBMCs were isolated using gradient centrifugation and used for immediate analyses as well as also being cryopreserved for subsequent studies. Serum tubes were allowed to stand upright for 2 hours at room temperature. Serum was subsequently isolated by centrifugation at 2,000 X g for 10 minutes and stored at - 80°C for later analysis.

### Safety assessment

Results from physical examination, including vital signs and body temperature, pulse, and blood pressure was recorded at each time-point for vaccination. Adverse events (AEs) following vaccinations were recorded from each study participant at each visit and classified as mild, moderate, or severe. Severe adverse events were, when so deemed, classified as serious adverse events (SAEs). AEs and SAEs recorded for all study participants were deemed unlikely, possible, or probably related to the vaccinations.

### Determination of YFV RNA levels in serum

YFV-specific real-time PCR was used to determine the viral RNA in serum of study participants. RNA was isolated from 150 µL of serum using a NucleoSpin RNA Virus Kit (Machery-Nagel). One step real-time PCR was performed using TaqMan Fast Virus 1-Step Master Mix (Applied Biosystems), a FAM-TAMRA-labeled probe and primers (Fisher Biotechnology) according to the manufacturer’s instructions. The primers and probe were all specific for the highly conserved NS5 gene of YFV [40] and were used at a final concentration of 800 nM and 125 nM for the primers and probe, respectively. Amplifications were performed in 25 µL reactions using a QuantStudio 5 real-time PCR machine (Applied Biosystems) under the following thermal cycling conditions: 5 min at 50°C, 20 s at 95°C, 45 cycles of 15 s at 95°C, followed by 1 min at 60°C. RNA from the YFV 17D was calibrated in relation to a standard curve commercially available in the Techne qPCR test for YFV (Techne) and used for quantification. The limit of detection was 20 copies per 1 mL.

### Clinical chemistry

C-reactive protein (CRP), albumin (ALB), creatinine, bilirubin, aspartate transaminase (AST), alanine aminotransferase (ALT), alkaline phosphatase (ALP), and gamma-glutamyltransferase (GGT) were analyzed at the Karolinska University Hospital accredited clinical chemistry laboratory at the first three time-points (days 0, 7, and 14).

### Absolute cell counts

Absolute numbers of cells expressing CD45, CD3, CD4, and CD8 in peripheral blood were measured using BD Trucount™ tubes (BD Biosciences) according to the manufacturer’s instructions. Briefly, 50 µL of anticoagulated blood was added to Trucount™ tubes within three hours after extraction and thereafter fluorescently stained for CD45, CD3, CD4, and CD8 for 15 minutes at RT in the dark. Samples were then fixed with 1X BD FACS lysing solution before acquiring data on a BD Accuri flow cytometer (BD Biosciences).

### Multiplex measurement of cytokine secretion

GM-CSF, IFN-ɣ, IL-2, IL-4, IL-6, IL-8, IL-10, TNF, IFN-ɑ2, MIP-1β, IL-12, IL-15 and IL-18 were measured in serum, diluted 1:4, from the first four time-points (days 0, 7, 14, and 30) from each study participant using a pre-designed 8-plex assay with a custom-designed 5 single-plex assay Bio-Plex Pro Human Cytokine Assay (Bio-Rad), according to the manufacturer’s instructions.

### Antibody analyses

Assessment of virus-specific IgG levels and neutralizing antibody titers (nAbs) against TBEV, JEV and/or YFV was performed on day 0, day 30, and at the final time-point of the clinical trial (day 210 for cohort A and cohort C, day 60 for B, D, and E). TBEV-specific IgG antibodies were assessed using an anti-TBE virus ELISA “Vienna” (IgG) kit (Euroimmun) and JEV-specific IgG antibodies were assessed using an anti-JEV ELISA (IgG) kit (Euroimmun) according to the manufacturer’s instructions. Both assays included complete virus lysates as coating antigen. Antibody levels equal to, or greater than, 120 were considered seropositive for TBEV-specific IgG and levels equal to, or greater than, 20 were considered seropositive of JEV-specific IgG, both as determined by the ELISA kit manufacturer. TBEV-specific IgG levels ≥1,000 Vienna units were recorded as 1,000 Vienna units and likewise, JEV-specific IgG levels ≥200 RU/ml were recorded as 200 RU/ml.

Virus neutralization was measured by rapid fluorescent focus inhibition test (RFFIT) as previously described with modifications for TBEV, JEV and YFV [41]. Briefly, serum from days 0, 30 and the final time-point (day 210 for cohort A and cohort C, day 60 for B, D, and E) were heat-inactivated and tested in serial dilutions with fifty 50% focus-forming doses of either TBEV (*93-783* strain), JEV (*Nakayama* strain) or YFV (*Asibi* strain) in flat-bottom 96-well plates. Following a 90-minute incubation of serum and virus, 50,000 trypsinized BHK-21 cells were added to each well and the plates were incubated for 24 hours. Following incubation, cells and virus were discarded and then fixed in 80% acetone for 1 hour at -20° C. After fixation, virus infected cells were detected by staining with optimized dilutions of mouse anti-TBEV (clone 1004134, Bio-Techne), mouse anti-JEV (clone 6B4A-10, Merck) or mouse anti-YFV (clone 2D12-A, Merck) followed by detection with Alexa Fluor 488 AffiniPure goat anti-mouse IgG (H+L) (Jackson ImmunoResearch). Virus infected cells were detected using fluorescence microscopy and an IRIS FluoroSpot reader (Mabtech). In each well, 20 fields were examined for fluorescent foci (infected cells) and fields with one or more foci were recorded. Titers are defined as 50% effective dose (ED50) and reciprocal titers of ED50 ≥5 were deemed positive. Titers less than 5 showed no neutralizing capacity or less than 50% reduction.

### Flow cytometry

Freshly isolated PBMCs were used for phenotypical analysis using the following antibodies: Live/Dead cell marker Near IR (Life Sciences), anti-CD19 (clone SIJ25C1) BUV 395, anti-CD4 (clone SK3) BUV 737, anti-CD16 (clone 3GB) Pacific Blue, anti-CD14 (clone MφP9) AmCyan, anti-Ki67 (clone B56) AF700 (BD Biosciences), anti-CD20 (clone 2H7) FITC, anti-CD123 (clone 6H6) AmCyan, anti-CD27 (clone O323) BV650, anti-CD38 (clone HIT2) BV785, anti-IgD (clone IA6-2) PE-Cy7, anti-IgG (clone HP6017) PE (BioLegend), anti-CD56 (clone N901) ECD, anti-CD3 (clone UCHt1) PE-Cy5 (Beckman Coulter), anti-CD8 (clone 3B5) Qdot 605 (Invitrogen) and anti-IgA (clone IS11-8E10) APC (Miltenyi Biotech). Cells were incubated with 50 µL of surface staining antibody mix diluted in PBS for 30 minutes at 4°C in the dark. Following incubation, the cells were washed twice in FACS buffer (2% FCS and 2 mM EDTA in PBS) and then fixed and permeabilized using Foxp3/Transcription Factor Staining kit (eBioscience) for 30 minutes at 4°C. Cells were then washed twice in permeabilization buffer (eBioscience) followed by intracellular staining. Antibodies against Ki67 and IgG in permeabilization buffer were added to the cells and incubated in the dark for 30 minutes at 4°C. Cells were finally washed with FACS buffer and data acquired on a BD LSR Fortessa (BD Bioscience). Analysis of acquired data was done with FlowJo software version 10 (FlowJo Inc).

### CXCL13 ELISA

Undiluted serum was thawed at room temperature and analyzed using a Quantikine Human CXCL13/BLC/BCA-1 ELISA (R&D Systems) according to the manufacturer’s instructions. Limit of detection of the assay ranged between 7.8 and 500 pg/mL.

### Statistics

Statistical analyses were performed using GraphPad Prism (GraphPad Software). Data sets were analyzed using non-parametric Wilcoxon matched-pairs signed rank test, Kruskal-Wallis, or Friedman tests. Dunn’s multiple comparisons test was used to correct for multiple comparisons where applicable. Handling of missing values due to missed visits (13 out of 1,150 visits) were determined before analysis according to the study protocol. In line with this, missing values were imputed in Figs using median values from the specific time-point in each cohort. All *p*-values <0.05 were considered to be statistically significant (\**p* < 0.05; \*\**p* < 0.01; and \*\*\**p* < 0.001).

## Results

### Study participants and vaccination schedule

140 study subjects were planned for the present clinical trial. Over the course of 17 months, 161 volunteers were screened, 145 of whom were found to be eligible for the clinical trial and enrolled into one of the seven cohorts (A1, A2, B1, B2, C, D, and E). All but six study participants completed the trial, leaving the clinical trial with a final 139 study participants (**Fig 1A**). These 139 study participants all completed the full duration of the study and were included in subsequent safety and immunogenicity assessments. Cohorts A1 and A2 (n=40) received the TBEV and YFV vaccine at day 0, and subsequent TBEV vaccine doses 1 month after the first dose, and at 5 months after the second dose. Cohort A1 (n=20) received TBEV and YFV vaccines in different upper arms and cohort A2 (n=20) in the same upper arm. Cohorts B1 and B2 (n=40) receive received the JEV and YFV vaccine at day 0, and subsequent JEV vaccine dose at 1 month after the first dose. Cohort B1 (n=20) received JEV and YFV vaccines in different upper arms and cohort B2 (n=20) in the same upper arm. Cohorts C (n=19), D (n=20) and E (n=20) received TBEV, JEV, or YFV vaccinations, respectively, following the same dose schedule as described in cohorts A and B (**Fig 1B**). The study participants were age and sex-matched across the cohorts (**Table 1**).

**Table 1.** Cohort characteristics

|  | **A1**<br>**(n=20)** | **A2**<br>**(n=20)** | **B1**<br>**(n=20)** | **B2**<br>**(n=20)** | **C**<br>**(n=19)** | **D**<br>**(n=20)** | **E**<br>**(n=20)** | **All cohorts**<br>**(n=139)** |
| --- | --- | --- | --- | --- | --- | --- | --- | --- |
| **Age (years)** |  |  |  |  |  |  |  |  |
| Average (SD) | 31.3 (9) | 28.9 (7.1) | 28.3 (10.2) | 27.8 (4.6) | 24.4 (5.9) | 26 (6.3) | 29.7 (9.5) | 28.1 (7.9) |
| Median (IQR) | 29 (10.5) | 25.5 (7.8) | 25 (6) | 27.5 (7) | 23 (4) | 25 (2.5) | 28.5 (11.8) | 26 (7) |
| Min, Max | 18. 53 | 22. 46 | 21. 53 | 22. 38 | 20. 46 | 18. 47 | 18. 54 | 18. 54 |
| **Gender (%)** |  |  |  |  |  |  |  |  |
| Male | 7 (35) | 6 (30) | 9 (45) | 10 (50) | 6 (31.6) | 7 (35) | 8 (40) | 53 (38.1) |
| Female | 13 (65) | 14 (70) | 11 (55) | 10 (50) | 13 (68.4) | 13 (65) | 12 (60) | 86 (61.9) |

### Adverse events

69.8% of the 139 included study participants reported one or more AEs during the course of the clinical trial (**Table 2**). 77.4% of all AEs across all cohorts were mild, 20.8% were moderate, and 1.8% were severe. 65.2% of all AEs across all cohorts were deemed unlikely related to vaccination, while 11.8% were deemed possibly related to vaccination and 23.1% were deemed probably related to vaccination (**Table 2**).

**Table 2.** Summary of registered adverse events

|  | **A1**<br>(n=20) | **A2**<br>(n=20) | **B1**<br>(n=20) | **B2**<br>(n=20) | **C**<br>(n=19) | **D**<br>(n=20) | **E**<br>(n=20) | **Total**<br>(n=139) |
| --- | --- | --- | --- | --- | --- | --- | --- | --- |
| **Total AE** | 40 | 51 | 25 | 19 | 36 | 31 | 19 | **221** |
| **Donors with at least one AE** | 18 | 15 | 12 | 10 | 15 | 16 | 11 | **97** |
| **Severity:** |  |  |  |  |  |  |  |  |
| Mild | 35 | 35 | 21 | 12 | 26 | 26 | 16 | **171** |
| Moderate | 4 | 15 | 4 | 7 | 9 | 5 | 2 | **46** |
| Severe | 1 | 1 | 0 | 0 | 1 | 0 | 1 | **4** |
| **Related to vaccination** |  |  |  |  |  |  |  |  |
| Unlikely | 25 | 36 | 18 | 8 | 21 | 20 | 16 | **144** |
| Possible | 5 | 3 | 2 | 8 | 5 | 1 | 2 | **26** |
| Probable | 10 | 12 | 5 | 3 | 10 | 10 | 1 | **51** |

The most frequent AEs were common cold-like symptoms followed by redness/tenderness at the injection site, headache, fever, and influenza-like symptoms. The latter four symptoms represent classical reactogenicity related responses.

Cohorts A1, A2 and C, all of which received TBEV vaccinations, had a greater number of total registered AEs compared to the other cohorts, ascribed to multiple (n=3) TBEV vaccinations. Cohorts B1, B2, and D, all of which received JEV vaccinations, had greater number of total registered AEs compared to the E cohort, concordantly ascribed to multiple (n=2) JEV vaccinations. Four AEs were deemed severe, all of which were determined SAEs. These SAEs included one case with hip dysplasia in a newborn child of a study participant, one case with pneumonitis, one case with depression, and one case with a fall, all four of which were determined to be unrelated to vaccination. A full list of the cohorts’ AEs is shown in **STables 1 and 2**.

### Viral RNA levels, clinical chemistry, and immunology

The YFV vaccine causes a mild infection, often with detectable virus in circulation. As a safety-related assessment, we measured serum YFV NS5-RNA levels in all cohorts vaccinated with the YFV vaccine in order to assess if concomitant vaccination had any effects on YFV viral replication. Across all study cohorts, most study participants receiving the YFV vaccine had detectable levels of YFV NS5-RNA seven days after vaccination (**Fig 2A**). In all but four study participants, no YFV NS5-RNA was detectable at 14 days following vaccination. Overall, no significant differences were observed between the study cohorts with the exception of cohort B2 which was found to have somewhat higher median levels of YFV NS5-RNA copies in serum (**Fig 2B**). Based on these findings, we conclude that concomitant vaccination did not generally affect YFV replication in the study participants receiving concomitant vaccination.

**Fig. 2.**
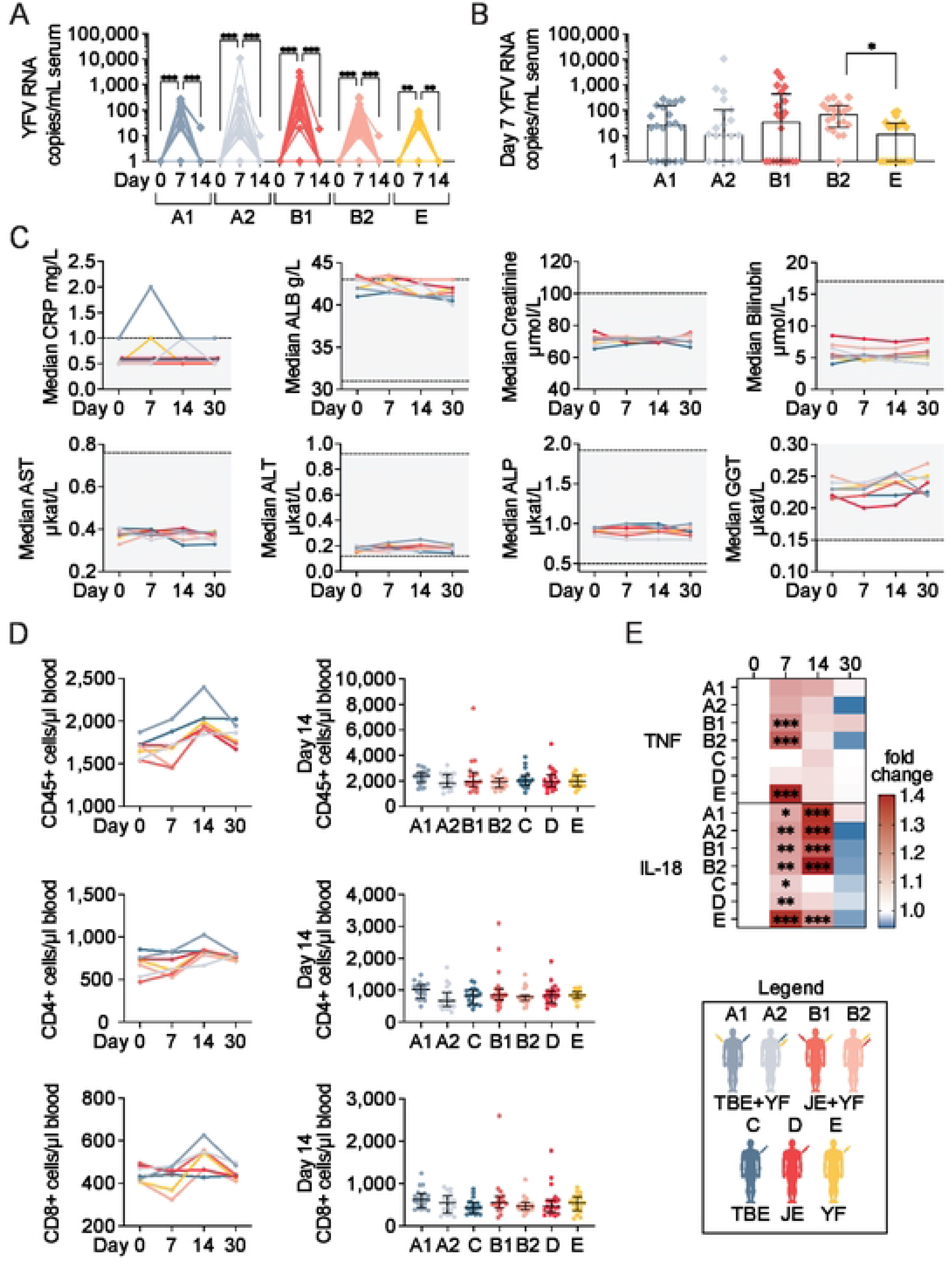
Clinical laboratory assessment following concomitant flavivirus vaccination. (**A**) Total number of YFV RNA copies/mL serum in cohorts receiving YFV vaccine. **(B)** Comparison of day 7 YFV RNA copies/mL between cohorts (plot depicts median with IQR). **(C)** Median clinical chemistry data-points during first four time-points of the study. Grey backgrounds denote the normal range in healthy adults. **(D**) Total numbers of CD45^+^, CD4^+^, and CD8^+^ cells over time following vaccination (left panels) and comparison of day 14 median absolute cell numbers in the respective vaccine cohorts (right panels, plots depict median with IQR). **(E)** Heat-map of fold change of serum TNF and IL-18 levels following the first vaccine dose. The legend denotes color-coding of the seven cohorts. Statistical analyses in **(A)**, **(C)**, and **(E)** were performed using nonparametric Friedman test with Dunn’s multiple comparison tests, in **(B)** and **(D** day 14 plots), using nonparametric Kruskal-Wallis test with Dunn’s multiple comparison tests, and in **(D** timeline) using nonparametric Wilcoxon matched-pairs signed-rank tests. *p < 0.05, **p < 0.01, ***p < 0.001.

As YFV infection can lead to liver and kidney damage, we assessed common inflammation as well as kidney- and liver-related clinical chemistry analytes to evaluate the effects of concomitant vaccination. No significant changes were observed over time in the cohorts with most clinical chemistry values lying within the normal range for healthy individuals (**Fig 2C**). These findings suggest that TBEV or JEV vaccination together with the YFV vaccine have no major systemic effects on kidney or liver organ function.

Further laboratory assessment included basic cellular immunological measurements following concomitant vaccination. This included standardized clinical diagnostic TruCount assays, in which absolute numbers of CD45^+^, CD4^+^, and CD8^+^ cells were measured (**Fig 2D**). Increases in CD45^+^ and CD4^+^ cell numbers were, as expected, observed at day 14 after vaccination in all cohorts. Similarly, CD8^+^ cells increased in absolute number 14 days following vaccination, however, only in cohorts receiving the YFV vaccine. There were no general differences in absolute cell counts at day 14 between the study cohorts when assessing overall patterns compared to days 0, 7 and 30, respectively.

As a final laboratory measurement assessing concomitant vaccination, multiple soluble cytokines and chemokines were measured in serum before and at the three first time points following vaccination in all cohorts. Of these, IL-18, IL-8, TNF and MIP-1β were at detectable levels among most study participants while GM-CSF, IFN-ɣ, IL-2, IL-4, IL-6, IL-10, IFN-ɑ2, IL-12, and IL-15 were below detection levels in the majority of study participants (**S1 Fig**). Notable increases in TNF and IL-18 levels were observed in all cohorts receiving the YFV vaccine (cohorts A1, A2, B1, B2, and E) (**Fig 2E**). There were no statistically significant differences in TNF or IL-18 levels between these cohorts at any of the measured time-points.

Taken together, no marked clinical safety-related effects following concomitant vaccination of the TBEV or JEV vaccines with the YFV vaccine were observed. Nor were there any statistical differences observed when vaccines were administered in the same or different upper arms of study participants.

### Virus specific IgG levels

As part of the primary endpoint, TBEV and JEV specific IgG levels were measured before vaccination and at days 30 (30 days after dose 1) and 210 (30 days after dose 3) for the TBEV vaccine cohorts, and at days 30 (30 days after dose 1) and 60 (30 days after dose 2) for the JEV vaccine cohorts.

Study participants from cohorts A1, A2, and C all developed IgG antibodies against TBEV by the end of the trial (**Fig 3A**). A total of six study participants from these three cohorts had pre-existing antibodies at day 0 by using the detection threshold (≥ 120 Vienna units) for positive response as defined by the manufacturer. For all cohorts, a gradual increase of virus specific IgG levels was observed as assessed at day 30 and day 210 (**Fig 3A**). All responses were above the positive threshold at the final time-point (**Fig 3B**). Cohorts A1, A2, and C did not differ significantly from each other (**Fig 3B**). The majority of study participants from cohorts B1, B2, and D developed IgG antibodies against JEV by the end of the trial (**Fig 3C**). A total of 18 study participants from these three cohorts presented with pre-existing antibodies at day 0 according to the manufacturer’s positive threshold (≥ 20 RU/mL). For all cohorts, a gradual increase of virus specific IgG levels was observed as assessed at day 30 and day 60 (**Fig 3C**). At the final time-point, no significant differences in antibody titers were observed between cohorts B1, B2, and D (**Fig 3D**). Consequently, cohorts B1 and B2 did not differ significantly from each other (**Fig 3D**).

**Fig. 3.**
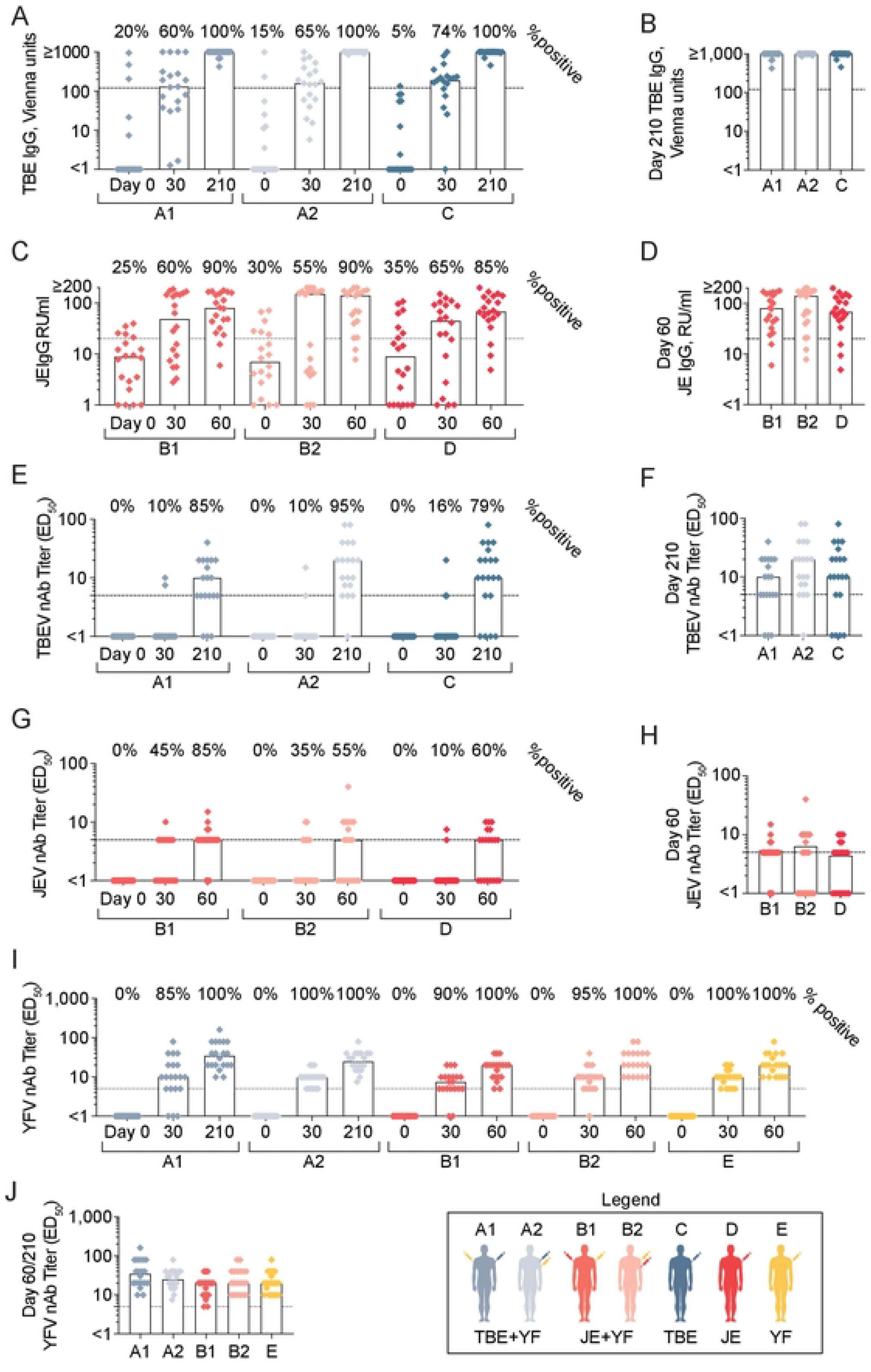
Seroconversion and development of neutralizing antibody following concomitant flavivirus vaccination. (**A**) TBEV specific IgG levels before and after concomitant vaccination with TBEV and YFV or only TBEV. The dotted line denotes manufacturer’s positive threshold and samples ≥1000 Vienna units were plotted at 1000 Vienna units. **(B)** Comparisons of final TBEV-specific IgG levels between cohorts A1, A2 and C. **(C)** JEV specific IgG levels before and after concomitant vaccination with JEV and YFV or only JEV. The dotted line denotes manufacturer’s positive threshold and samples ≥200 RU/mL were plotted at 200 RU/mL. **(D)** Comparisons of final JEV-specific IgG levels between cohorts B1, B2 and D. **(E)** TBEV neutralizing antibody titers before and at days 30 and 210 after concomitant vaccination with TBEV and YFV or only TBEV. Study participants with nAbs detected at day 0 (A1, n =1; A2, n = 1) excluded from figures. The dotted line denotes positive threshold (≥ 5). **(F)** Comparisons of final nAb titers against TBEV between cohorts (percent seropositive individuals as denoted above). **(G)** JEV neutralizing antibody titers before and at days 30 and 60 after concomitant vaccination against JEV and YFV or only JEV. Dotted line denotes positive threshold (≥ 5). **(H)** Comparisons of final nAbs titers against JEV between cohorts (percent seropositive individuals as denoted above). **(I)** YFV neutralizing antibody titers before and at days 30 and 60 after concomitant vaccination with TBEV or JEV and YFV, or only YFV. The dotted line denotes positive threshold (≥ 5). **(J)** Comparisons of final nAbs titers against YFV between respective cohorts. The legend denotes color coding of the seven cohorts. All plots are depicted with median values. Statistical analyses in **(A), (C), (E), (G),** and **(I)** were performed using nonparametric Friedman test with Dunn’s multiple comparison tests and in **(B), (D), (F), (H),** and **(J)** using nonparametric Kruskal-Wallis test with Dunn’s multiple comparison tests. *p < 0.05, **p < 0.01, ***p < 0.001.

### Virus neutralization

Apart from the assessment of antigen-binding IgG responses, rapid fluorescence focus inhibition tests (RFFIT) were performed to measure nAb titers against TBEV, JEV, and YFV to compare the effect of concomitant vaccinations with respective single vaccination outcomes. All study participants were screened at day 0 for nAbs against the three flaviviruses. Two study participants had nAbs against TBEV at day 0 (one each from cohorts A1 and A2, respectively). These were excluded from subsequent analyses. No study participants had preexisting nAbs against JEV or YFV at day 0.

Few study participants developed TBEV nAbs at day 30, however, the majority developed positive titers by day 210 (30 days after third dose) (A1: 85%, A2: 95%, and C: 79%) (**Fig 3E**). There were no statistically significant differences in nAb titers between cohorts A1, A2, and C (**Fig 3F**). Similarly, there were no statistically significant differences in nAb titers when cohorts A1 and A2 were compared (**Fig 3F**). Similar to TBEV, few study participants developed JEV nAbs at day 30, however, the majority developed positive titers by day 60 (30 days after third dose) (B1: 85%, B2: 55%, and D: 60%) (**Fig 3G**). There were no statistically significant differences in nAb titers between cohorts B1, B2, and D (**Fig 3H**). There were no statistically significant differences in nAb titers when cohorts B1 and B2 were compared (**Fig 3H**). All study participants from the cohorts receiving YFV vaccination developed nAbs by day 60 (**Fig 3I**). Additionally, there were no statistically significant differences in YFV nAb titers between cohorts A1, A2, B1, B2, and E (**Fig 3J**).

No statistical differences were observed in terms of nAb titers were observed when concomitant vaccinated study participants had received their respective vaccine in the same upper arm (2.5 cm distance according to protocol) versus in different upper arms. A trend towards higher titers (mean value) of nAb was noted in the groups where vaccines were given in the same upper arm (cohorts A2 and B2) versus different upper arms (cohorts A1 and B1) (**Figs 3F and H**).

The design of the clinical trial also allowed us to divide the enrolled study subjects based on gender. No statistically significant differences were observed between males and females in terms of final nAb titers against TBEV, JEV, or YFV (**Fig S2**). Noteworthy, however, an indication of an increase in females versus males in the TBEV-specific nAb titers were observed when assessing all study participants receiving the TBEV vaccine (**Fig S2A, right panel**).

In summary, concomitant TBEV or JEV vaccination with YFV vaccine did not affect the development of virus-specific nAbs towards TBEV or JEV. Likewise, YFV nAb titers did not differ between the cohorts, whether the YFV vaccine was administered alone or together with the TBEV or JEV vaccines (**Fig 3J**).

### Activation of B cells, T cells and NK cells

In addition to the virus-specific binding IgG antibody responses and nAbs we, we expanded the analyses in exploratory studies to investigate specific cellular responses following vaccination of the different cohorts. Freshly isolated PBMCs from each time-point were used to assess the effects of concomitant vaccinations on lymphocyte activation. Co-expression of CD38 and Ki67 in B cells, CD4^+^ T cells, CD8^+^ T cells, and NK cells (CD56^dim^ population of NK cells when not else noted) was measured (**Fig 4A**). Cohorts A1, A2, B1, B2, and E (all vaccinated with the YFV vaccine) exhibited increased frequencies of activated B cells, CD4^+^ T cells, CD8^+^ T cells, and NK cells at day 14 following vaccination (**Fig 4B-E**). A trend towards earlier activation of NK cells was observed in some study participants (**Fig 4E**), in line with previous reports [42, 43]. In general, vaccination with only YFV vaccine or vaccination with concomitant vaccination with YFV vaccine and TBEV or JEV vaccine led to a stronger activation among these cellular subsets compared to single vaccination with the TBEV or JEV virus vaccine (**Fig 4B-E**). No statistically significant differences in lymphocyte activation were observed between cohorts A1 and A2, as well as between B1 and B2 (**Fig 4B-E**), with the exception of a more marked CD4^+^ T cell activation in group B2 versus group B1 (**Fig 4C**).

**Fig. 4.**
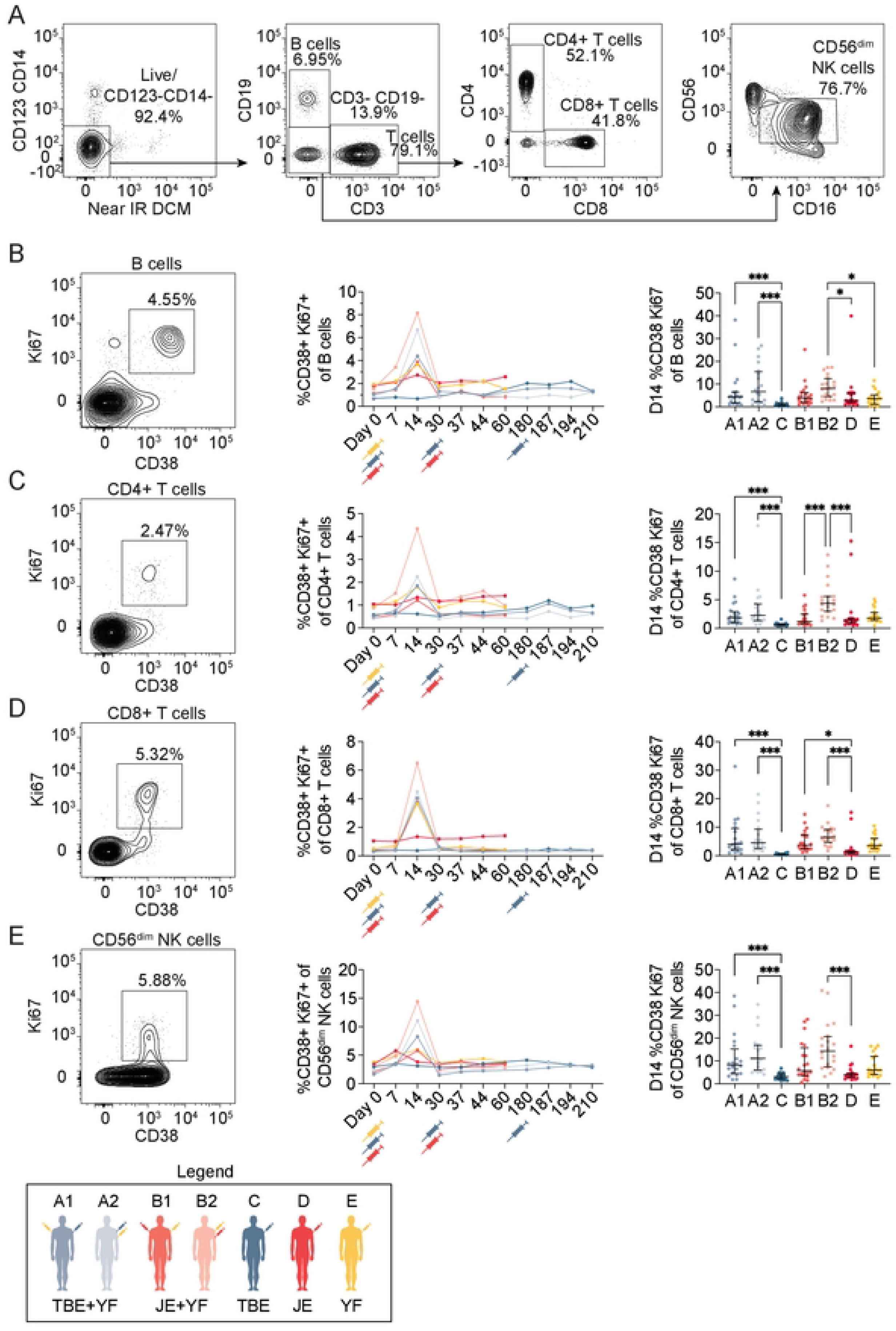
B cell, T cell and NK cell activation following concomitant flavivirus vaccination. **(A**) Representative gating strategy identifying B cells, CD4^+^ and CD8^+^ T cells as well as CD56^dim^ NK cells. Representative FACS plot of CD38^+^ Ki67^+^ **(B)** B cells, **(C)** CD4^+^ T cells, **(D)** CD8^+^ T cells, and **(E)** CD56^dim^ NK cells (left panels) with corresponding plot of the respective cohorts’ frequency of activation medians before and following vaccination program (middle panels). Plots comparing peak activation at day 14 between the vaccination cohorts (right panels depicting median values with IQR). The legend denotes color-coding of the seven cohorts. Statistical analyses were performed using nonparametric Kruskal-Wallis test with Dunn’s multiple comparison tests. *p < 0.05 and ***p < 0.001.

### Germinal center activation and plasmablast responses

The observed activation of lymphocyte subsets indicates induction of innate and adaptive immune responses, likely linked to the development of protective immunity (nAbs). We therefore further explored parameters related to induction of humoral immunity such as germinal center activity and plasmablast expansion following vaccination.

First, CXCL13 levels in serum were measured as a proxy to germinal center activity. Elevated levels of serum CXCL13 were observed at days 7 and/or 14 following each vaccination in all cohorts, with the most marked changes seen amongst vaccine cohorts receiving the YFV vaccine (cohorts A1, A2, B1, B2, and E) (**Fig 5A**). Plasmablast expansion as a result of germinal center formation was also analyzed. An increase in the number of plasmablasts was observed 14 days after vaccination in all cohorts receiving the YFV vaccine (cohorts A1, A2, B1, B2 and E) (**Fig 5B and C**). Similar to B cell activation (**Fig 4B**), the concomitant vaccine cohorts (cohorts A1, A2, B1, B2) had significantly higher frequencies of plasmablasts at day 14 than the single TBEV and JEV vaccine cohorts (cohorts C and D). Additional deeper analyses were also performed, including studies of plasmablast specific immunoglobulin expression over time (**S3 Fig**).

**Fig. 5.**
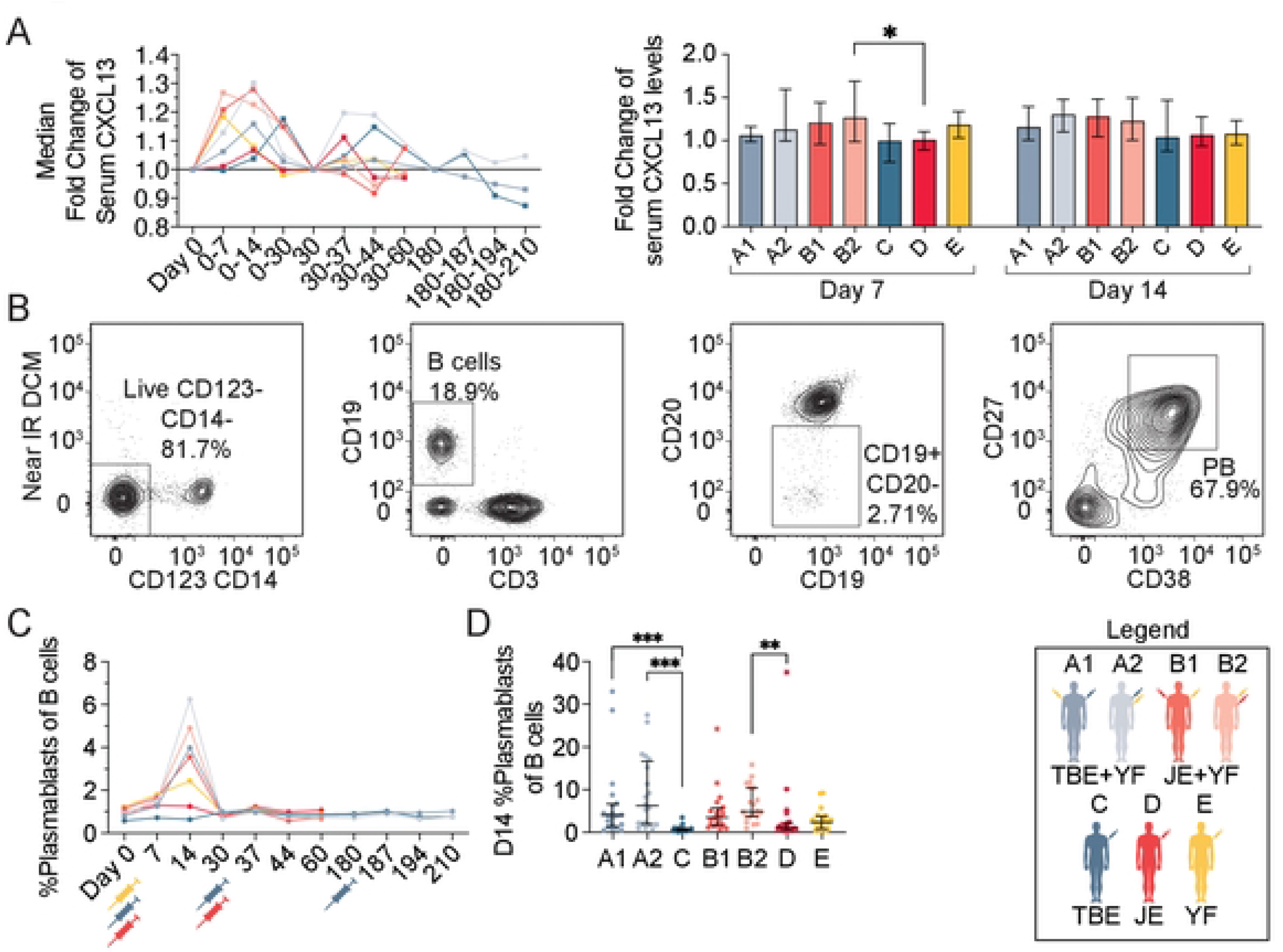
Germinal center activity and plasmablast expansion following concomitant flavivirus vaccination. **(A)** Median CXCL13 serum fold changes over time following vaccination program (left panel), and comparison of peak levels between cohorts at days 7 and 14 (right panel). **(B**) Representative gating strategy for identifying plasmablasts. **(C)** Median plasmablast expansion over time following vaccination and **(D)** comparison of peak expansion between cohorts at day 14. The legend denotes color-coding of the seven cohorts. All plots are depicted with median values with IQR. Statistical analyses were performed using nonparametric Kruskal-Wallis test with Dunn’s multiple comparison tests. *p < 0.05, **p < 0.01, ***p < 0.001.

Together these findings indicated that concomitant TBEV or JEV vaccination with the YFV vaccine leads to a marked germinal center activation and plasmablast expansion, likely resulting to a significant extent from the YFV vaccine response.

## Discussion

Here, we report the results of an open label, non-randomized, academic clinical trial assessing the safety and immunogenicity upon concomitant vaccination with different commonly used flavivirus vaccines. Healthy study participants were enrolled into cohorts receiving vaccines against TBEV and YFV in different or the same upper arm, JEV and YFV in different or the same upper arm, and in control cohorts receiving vaccines against TBEV, JEV, or YFV. Co-administration of the TBEV or JEV vaccine with the YFV vaccine was well tolerated with respect to reported AEs. Clinical virology, chemistry, and immunological assessments supported these conclusions with no markedly unexpected responses observed. Serological responses including virus-specific IgG levels and virus-specific nAb titers towards individual viruses were not adversely affected by concomitant vaccination. Additional immunological studies added deeper immunological insights into vaccine responses in concomitantly vaccinated study participants and controls.

Of the AEs reported following vaccination, the majority were mild, dominated by common cold symptoms followed by classical reactogenicity-related responses. The majority of the reported AEs were deemed unrelated to vaccination. AEs related to vaccination (deemed possible or probable) were in line with previous safety data of single vaccination studies and the manufacturer’s reported known side effects [15–17,44–46]. YFV RNA levels were generally similar between the concomitant and YFV only vaccine cohort, supported by normal clinical chemistry inflammation-, liver- and kidney-related parameters. Additionally, no marked differences were observed in absolute CD45^+^, CD4^+^, or CD8^+^ cell counts as well as TNF or IL-18 levels as well as other cytokine levels (data not shown), between concomitant and single vaccination cohorts.

Earlier studies have shown the possible enhancement of cross-reactive flavivirus-specific cellular and humoral immunity after flavivirus vaccinations [47–50]. These observations led us to address if concomitant vaccination of flavivirus vaccines had any deviating effect on the generation of virus-specific IgG and/or nAbs as well as on other immunological outcomes. All TBEV vaccinated study participants were well above the defined positive threshold and were found to have overall similar median IgG levels and nAb titers. In a similar manner, we assessed JEV-specific IgG and nAbs in the JEV vaccine cohorts. All but four study participants developed JEV-specific IgG levels above the positive threshold at the end of study with no differences between the concomitant vaccine cohorts and the single JEV vaccine cohort. Not all study participants receiving the JEV vaccine developed nAbs by the end of the study. Finally, all study participants receiving the YFV vaccine developed nAbs by the end of the study and no differences were found between the concomitant and YFV vaccine only cohorts. The findings suggest that concomitant vaccination had no general negative effect on the development of protective virus-specific serological responses.

In addition to assessing the effect of concomitant vaccination on serological outcomes, we also assessed effects on lymphocyte activation and, in more detail, germinal center and plasmablast responses. As expected, NK cell, T cell, and B cell activation was observed in the cohorts receiving the live attenuated YFV vaccine (cohorts A1, A2, B1, B2 and E) in line with previous research [42,43,51]. Cohorts C and D showed little to no increase in activation of these cell subsets throughout the study, not unexpected from the inactivated TBEV and JEV vaccines, respectively. Despite the lack of early cellular activation amongst cohorts C and D, this had no effect on the outcome of development of TBEV and JEV nAbs as assessed at the end of the study. Similar patterns were observed in plasmablast expansion, a B cell subset responsible for secreting large amounts of antibodies following infection or vaccination [52]. Plasmablasts expanded robustly in cohorts receiving the YFV vaccine (cohorts A1, A2, B1, B2 and E), a finding consistent with previous studies of YFV vaccinated individuals [42,43,51,53–58]. Plasmablast expansion was not observed in the TBEV or JEV only cohorts (cohorts C and D) upon primary vaccination. However, subsequent doses of TBEV or JEV vaccines led to detection of some plasmablast expansion. As plasmablasts can derive from previously generated memory B cells [59, 60], more expansions may be expected after several doses of TBEV or JEV vaccine. In summary, few differences in immunological responses were observed between concomitant flavivirus vaccination and single vaccination, much of the responses observed likely due to the YFV vaccine.

Co-administration of vaccines into the same or different upper arms allowed for comparative studies between these different strategies of administering two different vaccines. One advantage with administrating vaccines in the same upper arm (or at the same site in general) is that the other upper arm is not affected in terms of local reactogenicity responses. That may save the dominant arm from possible functional impairment over the first days following vaccination. As mentioned, no statistically significant differences between the study and control cohorts’ TNF or IL-18 serum comparing cohorts A1 with A2 or B1 with B2 were observed. Similarly, no statistical differences were found in terms of the development of virus-specific nAbs between the A1 and A2 or B1 and B2 cohorts as well as other B cell-related responses including activation and plasmablasts expansion. The only observed statistically significant difference noted between the same arm (A2 and B2) and different arm (A1 and B1) concomitant vaccine cohorts was the frequency of activated CD4^+^ T cells being higher in cohort B2 versus cohort B1. This said, TNF and IL-18, general lymphocyte activation, nAb titers, as well as percentages of plasmablasts, were if anything slightly higher in the study groups where concomitant vaccination had occurred in the same upper arm (A2 and B2) versus different upper arms (A1 and B1). We cannot exclude the possibilities that adjuvant-like effects imposed by, e.g., the live attenuated YFV vaccine or priming of immune cells in the same lymph node might contribute to the effects observed.

The relative strength of the present study is that data were collected from a *bona fide* prospective clinical trial with the scientific and regulatory rigor inherent to this design, independent study monitoring of data, both of which contributed to data quality, and additionally low drop-out rates and minimal risk of selection bias. An additional advantage was the ability to include and compare two different inactivated whole virus vaccines, the TBEV and JEV vaccines, respectively. The study contributes carefully evaluated clinical safety data in addition to standard clinical virology, clinical chemistry, and clinical immunology data. The number and frequency of study participant visits added to the strength of the study. With respect to limitations of this study, even later sampling time-points would have allowed assessment of sustainability of the developed nAbs and larger study groups could have given even more robust data. Another relative limitation of the study is the fact that it did not include screening for TBEV, JEV or YFV seropositivity prior to enrollment into the study. However, only two study participants with positive nAbs against TBEV and no one to JEV or YFV were identified.

Concomitant vaccine administration strategies have been used in many child vaccination programs [61, 62] as well with travelers on their way to endemic regions [25, 63]. Administration of multiple vaccines at the same time is convenient as it reduces the number of clinical visits, thus saving time and resources, and provides protection against multiple pathogens in a shorter time-span when an individual may be vulnerable to infection. In addition, most reports deem concomitant vaccinations safe and effective [25,29,64,65]. In this respect, the guidelines of the US Food and Drug Administration (FDA) regarding the YFV vaccine YF-Vax (Sanofi Pasteur) state that it can be administered with the that measles vaccine, diphtheria and tetanus toxoid and whole cell pertussis vaccine (DTP), Hepatitis A and B vaccines, meningococcal vaccine, and typhoid vaccine (www.fda.gov). Most regulatory authorities including the Swedish Medical Products Agency recommend concomitant delivery of many vaccines, including the YFV vaccine, at separate injection sites (www.lakemedelsverket.se/en). With respect to flavivirus vaccines, the FDA specifically notes that the potential interference between the YFV vaccine and JEV vaccines have not been established. In context of the COVID-19 pandemic, new vaccine technologies have been successfully implemented and are now being tested when administered concomitantly with seasonal influenza vaccines. The safety and immunogenicity data generated from this clinical trial can provide insights into concomitant vaccination strategies using different types of vaccine formulations that could aid in guiding new vaccination strategies of emerging and re-emerging viral infections.

In conclusion, this clinical trial showed that concomitant TBEV or JEV vaccination with the live attenuated YFV vaccine can be administered with little risk of severe adverse events and, additionally, no impairment on the development of nAbs to the respective viruses. Finally, the vaccines can readily be administered in the same upper arm.

## Data Availability

All relevant data are within the manuscript and its Supporting Information files.

## Acknowledgments

We thank all study participants for participating in the clinical trial, the Karolinska Trial Alliance staff and research nurses for organizing the recruitment and scheduling of study participants, vaccination, and clinical sampling. Additionally, we thank J. Verner-Carlsson and staff scientists at the Swedish Public Health Agency for support with neutralization assays and BSL-3 instruction. Further thanks to W. Christ for support in the BSL-3 facility, P. Jahnmatz for discussions around the FluoroSpot reader assessment, and D. Wullimann for discussions and advice on the project.

## Author Contributions

Conceptualization: KB and H-GL; data curation: JTS, ML, KB; formal analysis: JTS, ML, KB; funding acquisition: KB, H-GL; investigation: JTS, ML, RV, JE, KB; development of methodology: JTS, ML, KB, KL; management and coordination responsibility: JTS, ML, KB, H-GL; provision of study materials and samples: NA-T, LL, KB, H-GL; oversight and leadership: JK, KL, KB, H-GL; verification: JTS, ML, NA-T, JK, KB; data visualization: JTS, ML, KB, H-GL; writing initial draft: JTS, ML, KB, H-GL; manuscript review and revision: JTS, ML, RV, JE, NA-T, LL, SG-R, JK, KL, KB, H-GL.

## Abbreviations

AE: adverse event
Ab: antibody
ED50: 50% effective dose
ELISA: enzyme linked immunosorbent assay
JEV: japanese encephalitis virus
nAb: neutralizing antibody
PCR: polymerase chain reaction
RFFIT: rapid fluorescent focus inhibition test
SAE: serious adverse event
TBEV: tick-borne encephalitis virus
YFV: yellow fever virus

---

**S1 Fig.**
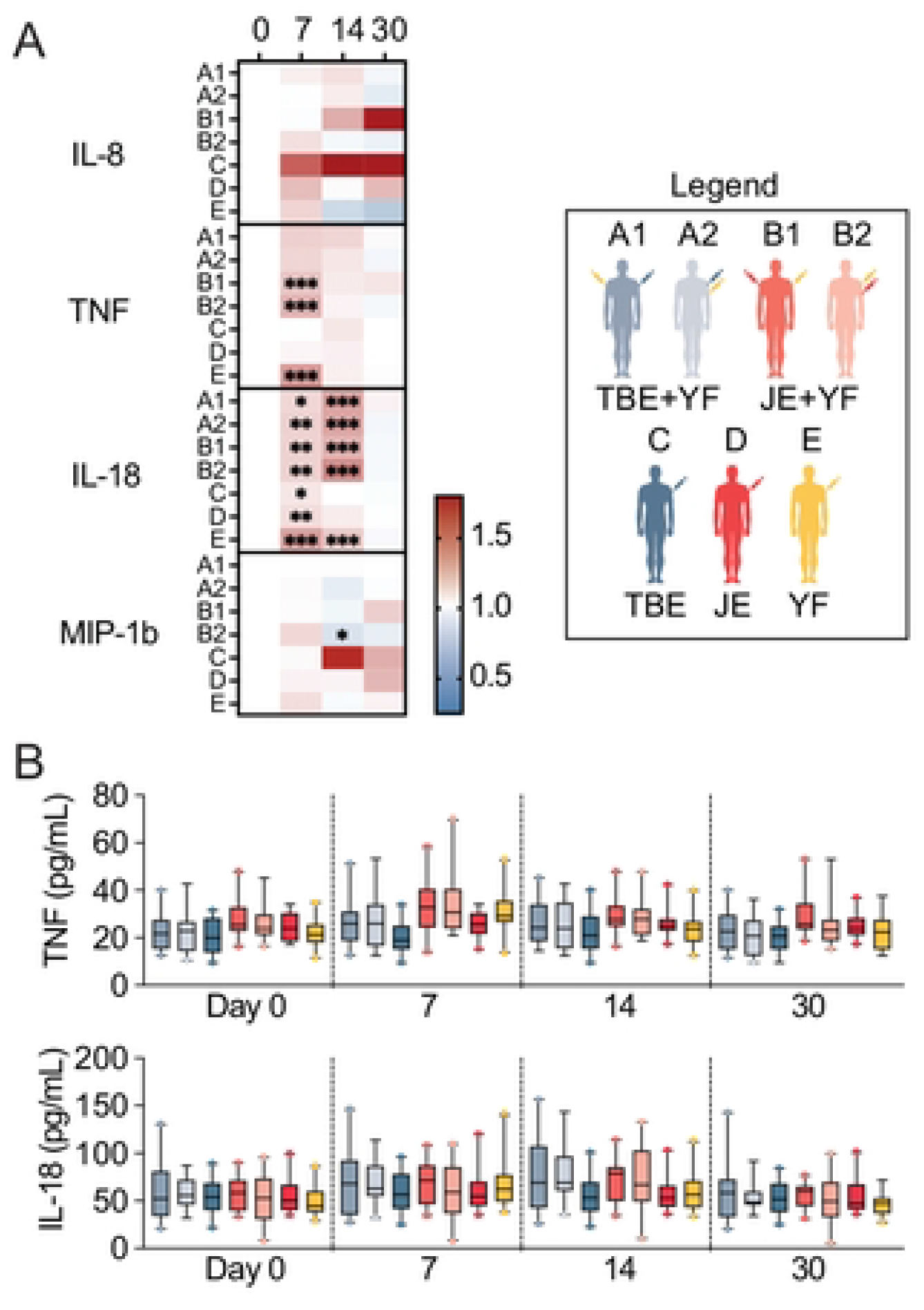
Soluble cytokine concentrations in serum following vaccination. (A) Heat map of fold changes of all detectable soluble factors in serum following vaccination. (B) Box and whisker plots comparing serum TNF and IL-18 concentrations following the first vaccine dose. The legend denotes color-coding of the seven cohorts. Statistical analyses were performed using nonparametric Friedman and Kruskal-Wallis test with Dunn’s multiple comparison tests.

**S2 Fig.**
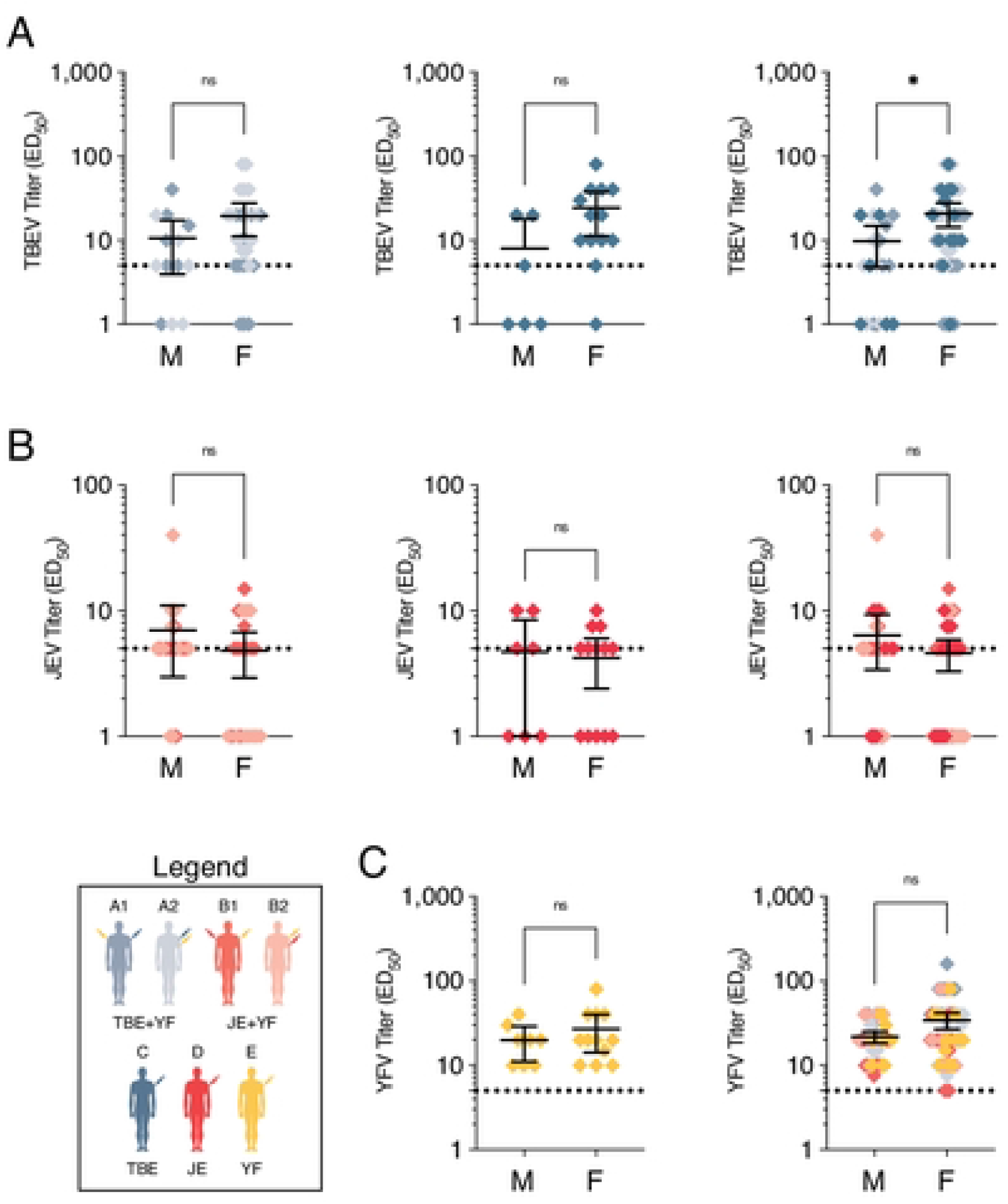
Sex-based comparisons of final virus nAb titers. (A) Comparisons of male (M) vs female (F) TBEV nAb titers at the final time point in cohort A1 and A2 (left panel), cohort C (middle panel) and combined A and C cohorts (right panel). (B) Comparisons of male (M) vs female (F) JEV nAb titers at the final time point in cohort B1 and B2 (left panel), cohort D (middle panel) and combined B and D cohorts (right panel). (C) Comparisons of male (M) vs female (F) YFV nAb titers at the final time point in cohort E (left panel) and combined A, B, and E cohorts (right panel). The legend denotes color-coding of the seven cohorts. Statistical analyses were performed using nonparametric Friedman and Kruskal-Wallis test with Dunn’s multiple comparison tests. *p < 0.05.

**S3 Fig.**
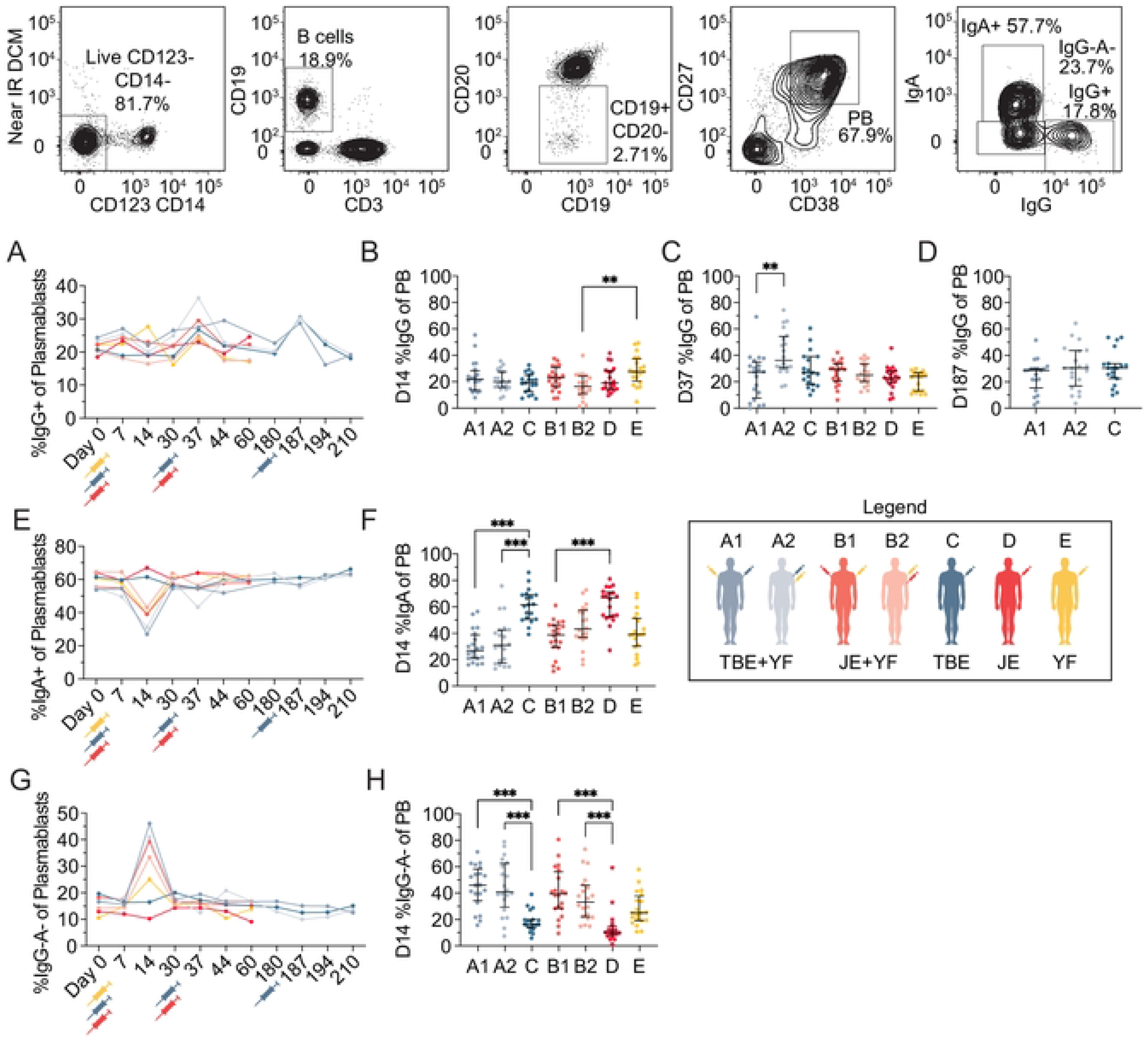
Immunoglobulin expression of circulating plasmablasts. (A) Median IgG expression of plasmablasts over time following vaccination and comparison of peak expression between cohorts at days (B) 14, (C) 37 and (D) 187. (E) Median IgA expression of plasmablasts over time following vaccination and (F) comparison of lowest expression between cohorts at day14. (G) Median IgG-A-expression of plasmablasts over time following vaccination and (H) comparison of peak expression between cohorts at day14. The legend denotes color-coding of the seven cohorts. All plots are depicted with median values with IQR. Statistical analyses were performed using nonparametric Kruskal-Wallis test with Dunn’s multiple comparison tests. *p < 0.05, **p < 0.01, ***p < 0.001.

**S1 Table.**
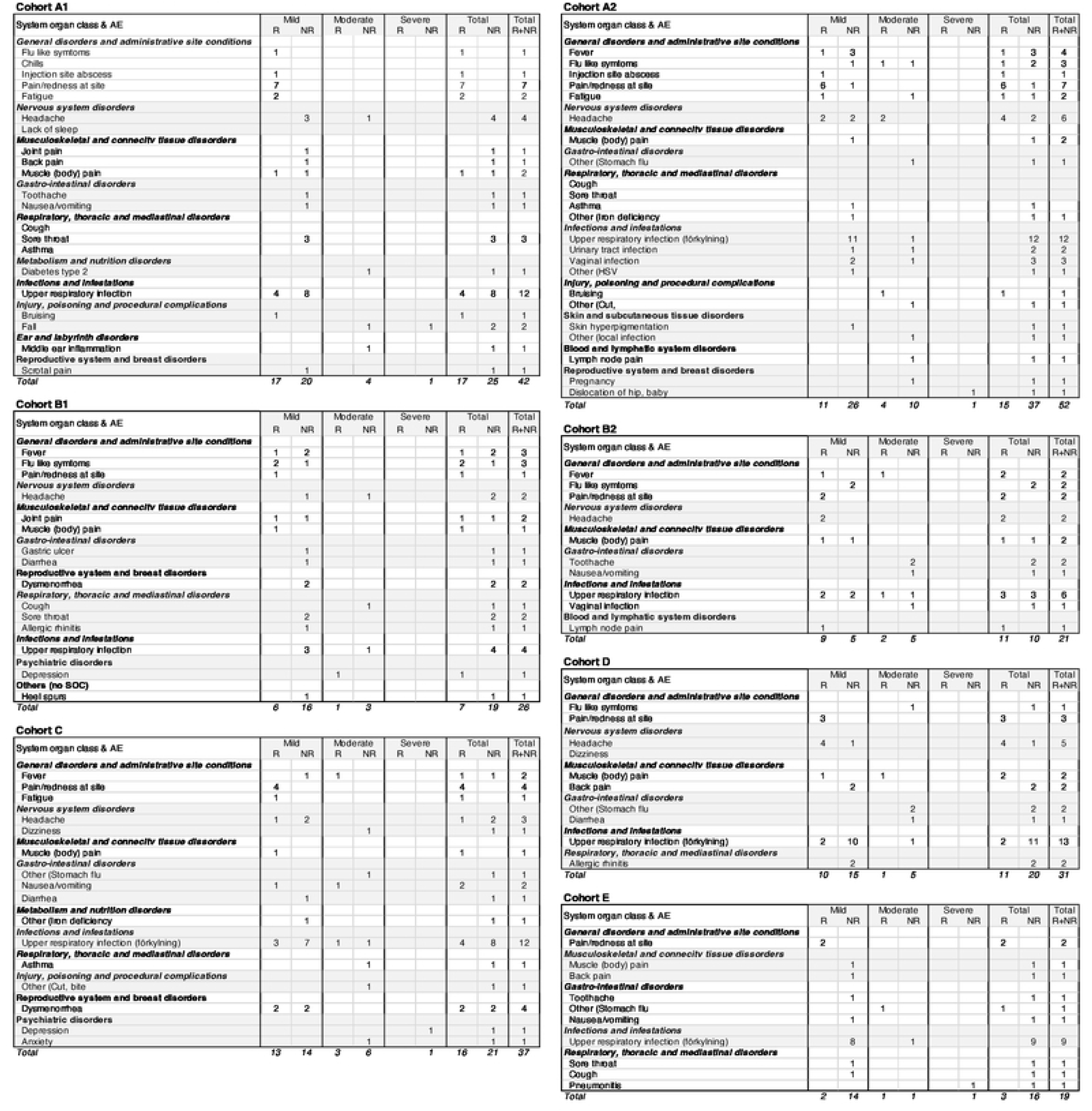
List of adverse events following vaccination

**S2 Table.** Summary of registered adverse events

|  | **A1**<br>(n=23) | **A2**<br>(n=20) | **B1**<br>(n=21) | **B2**<br>(n=21) | **C**<br>(n=20) | **D**<br>(n=20) | **E**<br>(n=20) | **Totalt**<br>(n=145) |
| --- | --- | --- | --- | --- | --- | --- | --- | --- |
| **Total AE** | 42 | 52 | 26 | 21 | 37 | 31 | 19 | **228** |
| **Donors with at least one AE** | 19 | 16 | 12 | 10 | 16 | 16 | 11 | **100** |
| **Severity:** |  |  |  |  |  |  |  |  |
| Mild | 37 | 36 | 22 | 14 | 27 | 25 | 16 | **177** |
| Moderate | 4 | 15 | 4 | 7 | 9 | 6 | 2 | **47** |
| Severe | 1 | 1 | 0 | 0 | 1 | 0 | 1 | **4** |
| **Related to vaccination** |  |  |  |  |  |  |  |  |
| Unlikely | 25 | 37 | 19 | 10 | 21 | 20 | 16 | **148** |
| Possible | 8 | 3 | 2 | 8 | 6 | 1 | 2 | **30** |
| Probable | 9 | 12 | 5 | 3 | 10 | 10 | 1 | **50** |
| **Related total** | **17** | **15** | **7** | **11** | **16** | **11** | **3** | **80** |

